# Time series modeling to estimate unrecorded burden of 12 symptomatic medical conditions among United States Medicare beneficiaries during the COVID-19 pandemic

**DOI:** 10.1101/2022.05.09.22274870

**Authors:** Michael Melgar, Jessica Leung, Jeffrey Colombe, Kathleen Dooling

## Abstract

**Objective:** U.S. healthcare utilization declined during the COVID-19 pandemic, potentially leading to spurious drops in disease incidence recorded in administrative healthcare datasets used for public health surveillance. We used time series modeling to characterize the magnitude and duration of the COVID-19 pandemic’s impact on claims-based monthly incidence of 12 symptomatic conditions among Medicare beneficiaries aged ≥65 years.

**Methods:** Time series of observed monthly incidence of each condition were generated using Medicare claims data from January 2016–May 2021. Incidence time series were decomposed through seasonal and trend decomposition using Loess, resulting in seasonal, trend, and remainder components. We fit a non-linear mixed effects model to remainder time series components and used it to estimate underlying incidence and number of unrecorded cases of each condition during the pandemic period.

**Results:** Observed incidence of all 12 conditions declined steeply in March 2020 with nadirs in April 2020, generally followed by return to pre-pandemic trends. The relative magnitude of the decrease varied by condition, but month of onset and duration did not. Estimated unrecorded cases during March 2020–May 2021 ranged from 9,543 (95% confidence interval [CI]: 854–15,703) for herpes zoster to 236,244 (95% CI: 188,583–292,369) for cataracts.

**Conclusions:** Due to reduced healthcare utilization during the COVID-19 pandemic, claims-based data underestimate incidence of non-COVID-19 conditions. Time series modeling can be used to quantify this underestimation, facilitating longitudinal analyses of disease incidence pre- and post-pandemic.

## Introduction

During the COVID-19 pandemic, there have been declines in utilization of all healthcare services in the United States, including elective and emergency visits.^1-8^ To limit transmission of SARS-CoV-2, social-distancing interventions and stay-at-home orders were implemented beginning with the national emergency declaration on March 13, 2020.^9^ In this context, an estimated 41% of adults had delayed or avoided care because of COVID-19 by June 2020.^2^ The health impact of delayed care can be significant, increasing the risk of delayed diagnosis and adverse outcomes; new cancer diagnoses in the Veterans’ Affairs healthcare system decreased by 13–23% during 2020 compared with pre-pandemic annual averages.^10^ Indeed, COVID-19-related healthcare avoidance and reduced access to healthcare likely contributed to all-cause excess mortality in 2020 and 2021, which far exceeded reported COVID-19 mortality.^11^

Administrative healthcare data are increasingly used in epidemiologic research, including disease surveillance and evaluations of population-level interventions such as health policy initiatives. Healthcare databases provide data on large populations at low cost.^12^ However, their detection of cases of disease depends on affected persons seeking medical care.^12^ Pandemic-associated changes in healthcare utilization may increase unrecorded cases of disease and therefore challenge longitudinal analyses of population-based incidence of non-COVID-19 conditions. To our knowledge, no work has been done to estimate the impact of the pandemic and social-distancing interventions on observed incidence of specific conditions using administrative healthcare databases. To measure this impact, we implemented a time series model of observed incidence of 12 symptomatic conditions among U.S. Medicare beneficiaries, characterized the magnitude and duration of pandemic-attributable declines in incidence following the national emergency declaration, and estimated the number of undetected cases of each condition during the pandemic period.

## Methods

We analyzed health claims during January 2016–May 2021 in fee-for-service claims data through the Centers for Medicare and Medicaid Services (CMS) Virtual Research Data Center, which includes a U.S. cohort of 50–60 million Medicare beneficiaries annually.^13^ Our study population consisted of beneficiaries aged ≥65 years with Medicare Parts A and B coverage, but not Part C (Medicare Advantage). Persons enrolled for only a portion of each calendar year were excluded from that year’s denominator. The CMS claims database is de-identified; therefore, upon review, this study was not considered human subjects research by the Centers for Disease Control and Prevention and did not require Institutional Review Board approval. This study was conducted in consistence with applicable federal law and CDC policy.^14^

We selected 12 symptomatic conditions that frequently trigger outpatient healthcare visits, are common among persons aged ≥65 years, and have been used in prior research to evaluate healthcare-seeking behavior:^15^ cataracts, cholelithiasis/cholecystitis, epistaxis, eyelid disorders (blepharitis, chalazion, hordeolum externum), gout, hemorrhoids, herpes zoster, ingrown nail, lipoma, nephrolithiasis, urinary tract infection, and venous thrombosis. An incident case of one of these conditions was identified as an individual’s first claim during the study period with a corresponding International Classification of Diseases, 9th or 10th Revision, Clinical Modification diagnostic code (Table 1).

**Table 1.** Diagnostic codes used to identify incident cases of 12 symptomatic conditions among enrollees in Medicare Parts A and B, but not C, January 2016□May 2021, United States.

| Condition | ICD-9-CM diagnostic codes | ICD-10-CM diagnostic codes |
| --- | --- | --- |
| **Cataracts** | 366.10, 366.14, 366.16, 366.17, 366.19 | H25.9, H25.04x, H25.1x, H25.8x |
| **Cholelithiasis and Cholecystitis** | 574.0x, 574.1x, 574.3x, 574.4x, 574.6x, 574.7x, 574.8x, 575.xx | K80.0x, K80.1x, K80.4x, K80.6x, K80.8x, K81.xx |
| **Epistaxis** | 784.7 | R04.0 |
| **Eyelid disorders** | 373.00, 373.2, 373.11 | H01.0x, H00.1x, H00.01x |
| **Gout** | 214.1, 214.8, 214.9 | D17.1, D17.2x, D17.3x, D17.7x, D17.9, M10.0x, M10.9 |
| **Hemorrhoids** | 455.0, 455.2, 455.3, 455.6 | K64.4, K64.8, K64.9 |
| **Herpes zoster** | 053.xx (excluding PHN codes, 053.12 and 053.13) | B02.xx (excluding PHN codes, B02.2x) |
| **Ingrown nail** | 703.0 | L60.0 |
| **Lipoma** | 214.1, 214.8, 214.9 | D17.1, D17.2x, D17.3x, D17.7x, D17.9 |
| **Nephrolithiasis** | 592.xx, 274.11 | N20.xx |
| **Urinary tract infection** | 599.0 | N39.0 |
| **Venous thrombosis** | 451.9, 453.8, 453.9 | I80.9, I82.6x, I82.8x, I82.9x |

Abbreviations: ICD-9-CM and ICD-10-CM, International Classification of Diseases, 9th and 10th Revisions, Clinical Modification, PHN, post-herpetic neuralgia

For each condition, we created a monthly time series of observed incidence per 1,000 person-years spanning the study period. Each time series was re-scaled by dividing by its mean value over the study period and was decomposed into seasonal, trend, and remainder components through seasonal and trend decomposition using Loess (STL) with robust fitting.^16^ Robust fitting was used to ensure that outlier values associated with reduced healthcare seeking behavior during the COVID-19 pandemic were captured in the remainder component, minimizing their impact on the seasonal and trend components. Loess windows for seasonal and trend extraction were 13 and 21 months, respectively.

The study period was divided into pre-pandemic (January 2016–February 2020) and pandemic (March 2020–May 2021) periods. Over the pandemic period, we fit a nonlinear mixed-effects model to the remainder time series components, modeling a decline in observed incidence as an alpha waveform function with rapid onset and a more gradual offset. At timepoint *t*_*j*_ the remainder component of the incidence time series of condition *I* was given by:

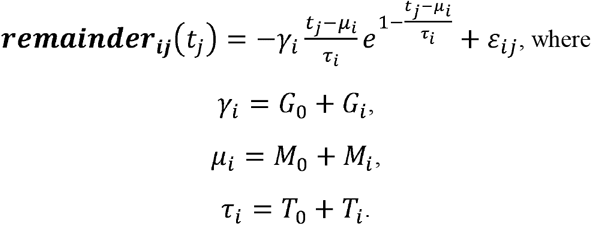

In the notation above, γ_*i*_ governed the relative magnitude of the fall in incidence of condition *i*, μ_*i*_ governed the onset of the fall (measured in months after February 2020, the last month before the national emergency declaration), and the time constant τ_*i*_ governed the duration of the fall (measured in months from onset to nadir). Each of these parameters was composed of a fixed effect (*G*_*0*_, *M*_*0*_, *T*_*0*_) and condition-specific random effects (*G*_*i*_, *M*_*i*_, *T*_*i*_). Twelve mixed-effects models were fit, each using data from all 12 conditions. The 12 models comprised every permutation of allowing and disallowing non-zero condition-specific random effects on each of the three parameters and, for models with non-zero random effects on more than one parameter, allowing and disallowing non-zero covariance between random effects (Table 2). Parameters were estimated by maximum likelihood using a quasi-Newton optimization method;^17^ optimization starting values were determined by first fitting an individual (fixed-effects) nonlinear model to each condition’s time series. Model goodness-of-fit was compared using the likelihood ratio test; p-values <0.05 were considered statistically significant. The model with the fewest degrees of freedom with goodness-of-fit not significantly inferior to that of another model was determined to be the most parsimonious.

**Table 2.**
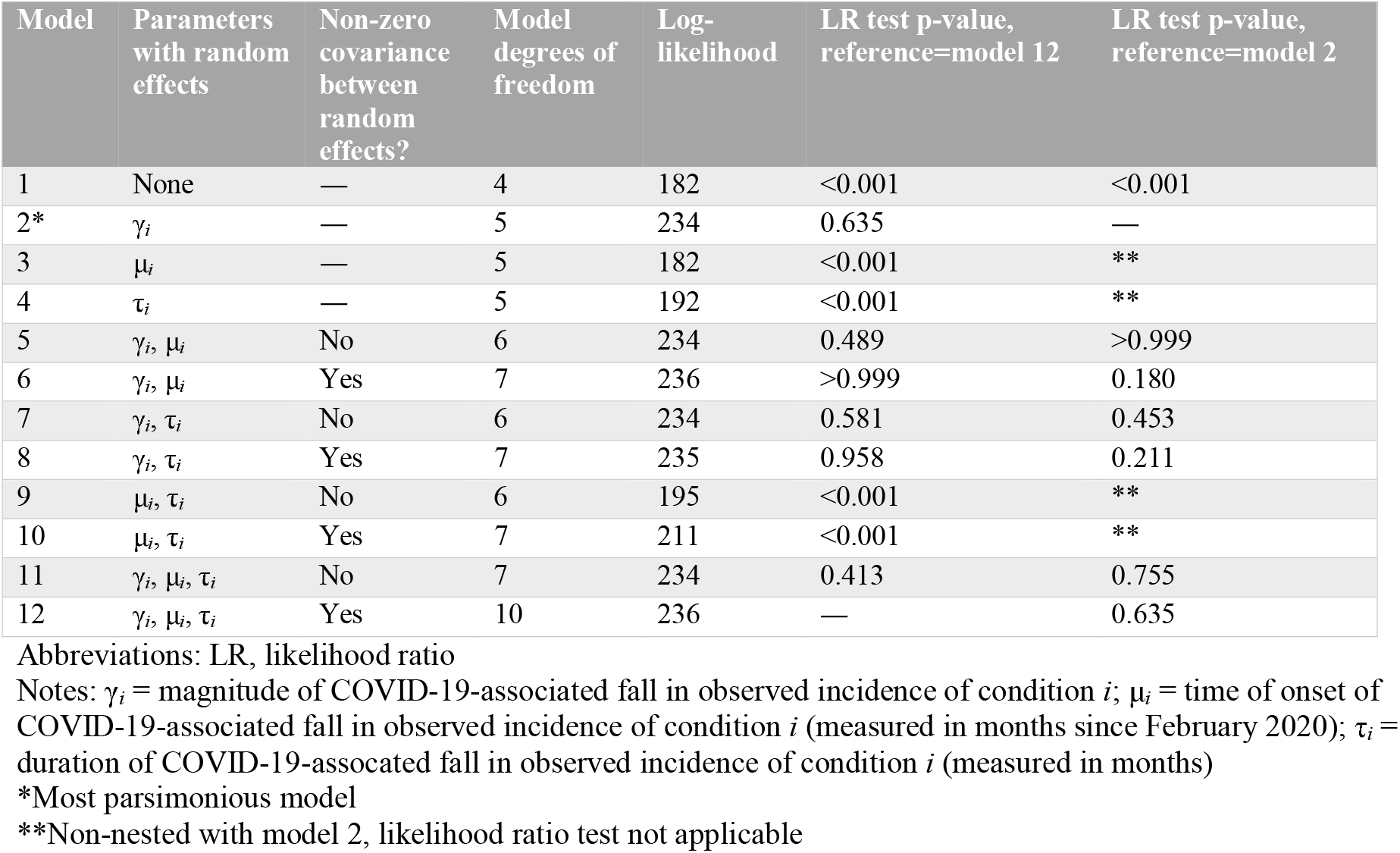
Goodness of fit of 12 nonlinear mixed-effects models of observed incidence of 12 symptomatic conditions among enrollees in Medicare Parts A and B, but not C, January 2016□May 2021, United States.

The most parsimonious model was used to estimate underlying incidence of each condition, including cases unrecorded due to pandemic-associated healthcare avoidance, by subtracting the alpha waveform from the model response function. We also estimated the total number of unobserved cases of each condition by calculating area-under-the-curve of the alpha waveform function during March 2020–May 2021. Ordinary bootstrap 95% confidence intervals (CI) were computed with 1,000 iterations.

Data preprocessing was performed using SAS 9.4 (SAS Institute, Cary, NC). Analysis was performed using R version 4.1.2 (R Core Team, Vienna, Austria).

## Results

The study population ranged from 25.7 million Medicare beneficiaries in 2020 to 26.1 million in 2019 (Table 3).

**Table 3.** Study population by year: enrollees in Medicare Parts A and B, but not C, January 2016□May 2021, United States.

| **Year** | **Study population** |
| --- | --- |
| 2016 | 25,929,309 |
| 2017 | 26,009,496 |
| 2018 | 26,010,775 |
| 2019 | 26,092,883 |
| 2020 | 25,704,617 |
| January–May 2021 | 25,818,561 |

**Table 4.** Number of unrecorded cases of 12 symptomatic conditions among approximately 26 million enrollees in Medicare Parts A and B, but not C, March 2020□May 2021, United States.

| Medical condition | Number of unrecorded cases (95% bootstrap confidence interval) |
| --- | --- |
| Cataracts | 236,244 (188,583–292,369) |
| Cholelithiasis/cholecystitis | 15,880 (10,410–21,332) |
| Epistaxis | 17,094 (11,068–23,234) |
| Eyelid disorders | 55,135 (39,856–71,427) |
| Gout | 24,747 (15,270–34,317) |
| Hemorrhoids | 113,850 (89,897–140,746) |
| Herpes zoster | 9,543 (854–15,703) |
| Ingrown nail | 39,668 (27,532–52,822) |
| Lipoma | 21,655 (16,972–27,424) |
| Nephrolithiasis | 34,709 (23,335–46,626) |
| Urinary tract infection | 72,656 (35,042–102,484) |
| Venous thrombosis | 10,394 (6,561–14,432) |

Of the 12 mixed-effects models of incidence, the most parsimonious included condition-specific random effects only on the relative magnitude of the fall in observed incidence, γ_*i*_ (Table 2, Model 2). The time of onset μ_*i*_ and time constant τ_*i*_ governing duration were fixed across conditions; allowing these parameters to vary by condition did not result in a significantly improved model fit.

Observed incidence of all 12 symptomatic conditions in the Medicare population exhibited unique seasonality and net decreasing trends over the study period (Figure). Seasonality of incidence of all conditions except epistaxis and nephrolithiasis exhibited annual nadirs in January. Epistaxis incidence exhibited annual peaks in winter months and nadirs in summer months. Nephrolithiasis incidence exhibited little seasonality, alternating higher and lower incidence months.

**Figure.**
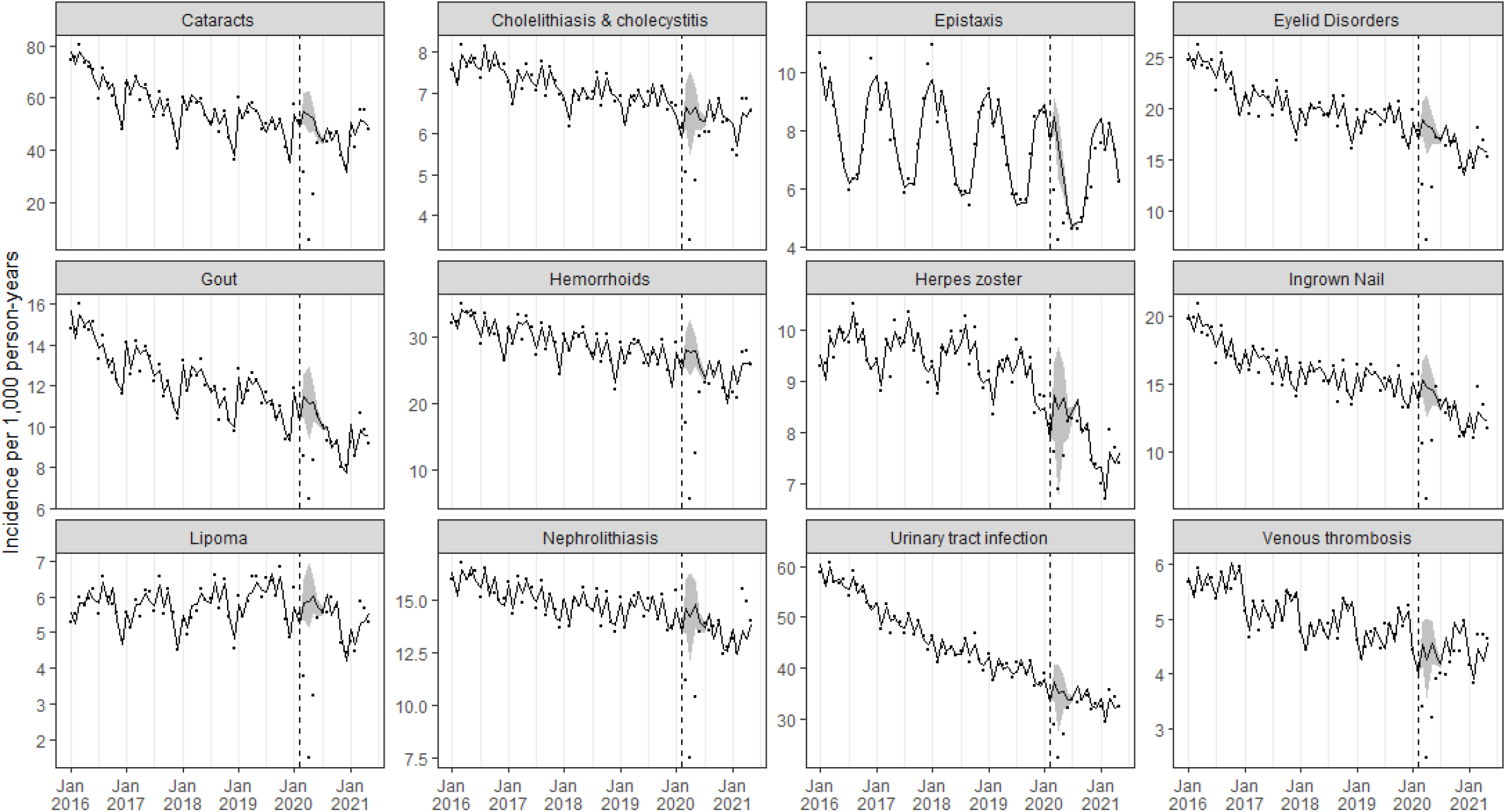
Trends in monthly observed incidence of 12 symptomatic conditions among approximately 26 million enrollees in Medicare Parts A and B, but not C, January 2016□May 2021, United States. Points indicate incidence recorded in the Centers for Medicare and Medicaid Services Virtual Research Data Center. Vertical dashed lines indicate February 2020 on the horizontal axis (the last month before the U.S. national COVID-19 emergency declaration). Solid black lines indicate estimated underlying incidence including cases unrecorded during the COVID-19 pandemic period. Shaded grey ribbons represent 95% bootstrap confidence intervals.

Incidence of all conditions exhibited sharp declines beginning in March 2020 with nadirs in April 2020 far below incidence expected from pre-pandemic seasonal and secular trends. However, by July 2020, observed incidence of most conditions approached pre-pandemic levels and trends. Model-estimated underlying incidence adhered closely to pre-pandemic trends throughout the pandemic period. Observed incidence of each condition fell outside of 95% CIs for estimated underlying incidence during March–May 2020.

There was high uncertainty in estimated underlying herpes zoster incidence during March–June 2020. After July 2020, herpes zoster incidence also declined more steeply than it did pre-pandemic, with a seasonal nadir in February 2021 below the April 2020 pandemic-associated nadir. Incidence of lipoma also appeared not to return to pre-pandemic levels by May 2021. Observed incidence of some conditions including herpes zoster, ingrown nail, and nephrolithiasis exhibited possible rebound effects in February–March 2021, exceeding model-estimated levels.

The estimated number of cases unrecorded due to pandemic-associated healthcare avoidance during March 2020–May 2021 ranged from 9,543 (95% CI: 854–15,703) for herpes zoster to 236,244 (95% CI: 188,583–292,369) for cataracts (Table 3).

## Discussion

Through time series modeling, we estimated the magnitude and duration of the effects of the early COVID-19 pandemic on claims-based observed incidence of 12 symptomatic conditions in a U.S. cohort of 26 million Medicare beneficiaries. All conditions in this analysis exhibited a simultaneous decline in observed incidence coinciding with the March 2020 national emergency declaration. Although the relative magnitude of the decrease varied by condition, the onset time and duration of the decrease were best modeled as constant across conditions, consistent with a common change in healthcare-seeking behavior. We estimated the unrecorded burden of disease during March 2020–May 2021 to be substantial: >10,000 cases for all conditions except herpes zoster and >100,000 cases for cataracts and hemorrhoids. Medically unattended cases of some conditions, such as venous thrombosis, may have contributed to excess mortality during the pandemic.^11^ Estimating this burden is critical to characterizing the pandemic’s full impact, including indirect consequences on health. This approach can be applied to other administrative datasets in longitudinal analyses of disease incidence spanning 2020–2022 and beyond. Accounting for the pandemic’s effect on healthcare-seeking behavior can address otherwise anomalous changes in observed incidence, facilitating impact measurement of other interventions of interest (e.g., cancer screening programs^18^, reimbursement^19^).

Our findings indicate a nadir in healthcare utilization in April 2020 and a subsequent return to pre-COVID trends, consistent with prior studies examining healthcare utilization.^1, 3, 10^ However, there was variability by condition. Monthly lipoma diagnoses appeared not to completely return to pre-pandemic baseline, possibly reflecting more persistent healthcare avoidance for a minimally symptomatic condition. Condition-specific analyses may better characterize incidence trends which may also have been impacted by factors other than COVID-19. For instance, herpes zoster incidence declined more steeply after May 2020 than it did pre-pandemic, with large uncertainty in incidence estimates during March–May 2020. Notably, recombinant zoster vaccine was licensed and recommended for U.S. adults aged ≥50 years in 2018 and may have had measurable impact on population zoster incidence.^20, 21^

One year into the pandemic, in March–April 2021, we found observed incidence of herpes zoster, ingrown nail, and nephrolithiasis to substantially exceed the model-estimated underlying incidence. This rebound effect may reflect undiagnosed burden of disease from earlier in the pandemic period. Patients may have managed associated symptoms without seeking medical care until later, when the perceived risk from COVID-19 was lower; spring of 2021 coincided with a fall in U.S. COVID-19 cases and hospitalizations relative to the 2020–2021 winter peak.^22^ Delayed healthcare seeking may result in poorer outcomes when patient ultimately present for care. It is unclear why a rebound effect was observed for only certain conditions and additional investigation, with perhaps a broader range of conditions may elucidate its cause.

This study has at least three limitations. First, we used diagnostic coding to identify incident cases and were unable to validate cases with medical record review. Second, incidence of the 12 conditions in persons aged ≥65 years may not be representative of incidence other acute conditions with differing severity or in other age groups, though our findings support those of other studies.^1, 3, 6, 10^ Finally, a control group unaffected by

COVID-19 was not available, so we could not rigorously attribute observed decreases in observed incidence to pandemic-associated effects. However, there was no analogous March–May drop in incidence during four years of pre-pandemic data for the same conditions in the same population, bolstering confidence that the changes were pandemic-associated.

Administrative healthcare data will continue to play a critical role in public health surveillance and other longitudinal epidemiologic studies. However, because the COVID-19 pandemic has challenged interpretation of observed disease incidence trends, analytic methods must address otherwise anomalous changes in incidence.

We propose a time series modeling approach to account for the effects of healthcare avoidance and reduced access on longitudinal disease incidence data that may facilitate continued surveillance using administrative data in the COVID-19 era.

## Data Availability

All data used in the present study are available from the Centers for Medicare and Medicaid Services Virtual Research Data Center.

https://resdac.org/

## Acknowledgements

We would like to acknowledge and thank Rafael Harpaz, MD, MPH for his work to identify the conditions included in this analysis.

